# Subthalamic Nucleus(STN) versus Globus Pallidus Internus(GPi) targeted with Deep Brain Stimulation in Parkinson’s Disease: A Systematic Review and Meta-Analysis of tremor outcome

**DOI:** 10.1101/2025.10.22.25338560

**Authors:** Farzan Fahim, MohammadAmin Farajzadeh, Seyyed Mohammad Hosseini Marvast, Afarinesh Hasheminejad, Maral Moafi, Ghazaleh Ghaffaripour Jahromi, Kimia Janeshin, Sayeh Oveisi, Alireza Zali

## Abstract

**Background:** Deep Brain Stimulation (DBS) is an established and effective treatment for patients with Parkinson’s disease (PD). Over recent decades, several brain targets have been explored for DBS, including the subthalamic nucleus (STN) and the globus pallidus internus (GPi). However, the relative efficacy of these targets in controlling tremor remains a subject of debate. This study aimed to systematically compare tremor outcomes between STN-DBS and GPi-DBS in patients with PD.

**Methods:** A prospectively registered protocol guided a comprehensive search of PubMed, Embase, Scopus and Web of science for randomized and controlled studies comparing tremor outcomes between STN-DBS and GPi-DBS. Standardized mean differences (Hedges’ g) in tremor reduction were extracted and synthesized using a random-effects model. The primary outcomes included the overall magnitude of tremor improvement, inter-study heterogeneity, and inter-individual variability in short-term response.

**Results:** Both STN-DBS and GPi-DBS yielded substantial and durable tremor reductions, typically ranging from 70% to 90% improvement from baseline. Meta-analytic pooling revealed no significant long-term difference between targets (Hedges’ g = –0.08; 95% CI, –0.53 to 0.38; p = 0.74), with minimal inter-study heterogeneity (I² = 0%). However, short-term postoperative data indicated a modest but consistent early advantage for STN-DBS in achieving faster tremor relief. Importantly, clinical follow-up findings revealed considerable variability in the magnitude and timing of tremor improvement across individual patients, independent of target.

**Conclusions:** STN-DBS and GPi-DBS are equally effective for long-term tremor control in PD. Nevertheless, the transient early benefit observed with STN stimulation and the pronounced individual variability in treatment response emphasize the importance of personalized DBS planning. Tailoring target selection and programming to patient-specific clinical profiles may optimize both short- and long-term outcomes

## Introduction

Parkinson’s disease is a common neurodegenerative disease that affects approximately 11.76 million people worldwide in 2021 and causes substantial disabilities, including motor, non-motor, and cognitive disabilities. Motor symptoms consist of tremor, muscle rigidity, stiffness, slowness, and imbalance, which will typically arise after loss of approximately half of the dopaminergic neurons in the caudal substantia nigra pars compacta ^1,2^.

Tremor occurs in about 75% of PD patients at disease onset and classically appears as a asymmetric resting tremor that can be enhanced by walking and performing tasks, such as calculation ^3^. In addition, it predominantly affects the upper limbs, and less frequently, the lower limbs, chin, jaw, or tongue ^4–7^. In comparison with other cardinal symptoms in PD patients, the underlying mechanism for the pathophysiology of tremor is unique and not fully understood, but recent advances in basic sciences and neurophysiology, particularly the introduction of new experimental techniques and the advent of functional magnetic resonance imaging (fMRI), have made it possible to clarify the mechanism ^8–10^. the effect of dopamine depletion in the appearance of tremor, the thalamus, including the VLpv is a key target regarding PD tremor, not the basal ganglia ^7,8,11^.

Deep brain stimulation (DBS) is a well-established surgical intervention for patients with advanced PD ^12,13^. it is a promising therapy, particularly in patients experiencing medication-resistant motor symptoms, motor fluctuations, or levodopa-induced dyskinesia (LID). DBS is more effective than best medical therapy in improving motor function and quality of life in well-selected PD patients ^14^.

Historically, surgical treatment for PD involved ablative procedures (lesioning)^15^. Since the 1950s, posterior part of the globus pallidus pars interna (GPi) was ablated. ^16^. With evolution into high stimulating techniques, ablating of STN as a key nuclous specially in animals, were showed improvement of parkinsonism Although STN lesions were initially associated with severe hyperkinetic or ballistic movement complications, by adjusting stimulator variables clinicians can control these side effects, making the STN a important target to reduce akinesia and rigidity ^17^.

Despite evidence showing that deep brain stimulation (DBS) of both the subthalamic nucleus (STN) and the globus pallidus internus (GPi) is effective for controlling Parkinson’s disease (PD), the important question is “which target provides superior tremor control?”. Historically, prospective but non-randomized studies sometimes suggested that STN stimulation was superior, effectively suppressing tremor, while GPi targeting reportedly had no significant effect on tremor ^18,19^. Conversely, other large studies and meta-analyses comparing overall motor benefit found no clear superiority for STN over GPi stimulation in the management of motor symptoms, including tremor^15,20,21^.

These persistent inconsistencies, data conflicts on motor functions, and timeline differences in achieving maximum benefit highlight the ambiguity in selection of best target to reduce tremor, underscoring the critical need for a comprehensive synthesis of the available evidence. Therefore, this systematic review aims to evaluate and compare the therapeutic effects of STN versus GPi targets in DBS, by focus on tremor, thereby clarifying this persistent and clinically gap.

## Methodology

This systematic review was performed based on Preferred Reporting Items for Systematic Reviews and Meta-Analysis (PRISMA) 2020 guidelines and its protocol was registered in the International Prospective Register of Systematic Reviews (PROSPERO, registration ID:CRD420251153336)

### Search strategy

A comprehensive literature search was conducted in PubMed, Scopus, Embase, and Web of Science (WOS) and Cochrane trials to identify all relevant studies, without any date or language restrictions. If any non-English studies were identified, their exact translation into English was planned. The search strategy was designed to capture studies comparing subthalamic nucleus (STN) and globus pallidus internus (GPi) deep brain stimulation (DBS) in Parkinson’s disease (PD).

For PubMed, the following search terms were used:

(“Parkinson Disease”[Mesh] OR parkinson*[tiab] OR PD[tiab])

AND (“Deep Brain Stimulation”[Mesh] OR “deep brain stimulation”[tiab] OR DBS[tiab])

AND ((“Subthalamic Nucleus”[Mesh] OR “subthalamic nucleus”[tiab] OR STN[tiab])

AND (“Globus Pallidus”[Mesh] OR “globus pallidus internus”[tiab] OR “internal globus pallidus”[tiab] OR GPi[tiab]))

AND (compar*[tiab] OR head-to-head[tiab] OR versus[tiab] OR vs[tiab] OR random*[tiab] OR trial[tiab])

Equivalent search strategies were adapted for Scopus, Embase, and Web of Science using their respective indexing terms and syntax. The detailed database-specific strategies are provided in Supplementary 1.

In total, 5926 studies were found: 2144 from Embase, 1538 from Web of Science, 1453 from Scopus, and 791 from PubMed. Duplicate records were removed using Covidence, which automatically identified 2641 duplicates, and an additional 80 duplicates were identified manually.

After removing duplicates, titles and abstracts were screened by (GG and AH and MH,MM), resulting in 112 articles selected for full-text review. To ensure screening reliability, two reviewers (GG and MH) independently assessed 20% of the full-text articles (n = 25). The distribution of their decisions was as follows: both excluded 21 studies, both included 3, and one case involved a minor disagreement (maybe vs exclude). The agreement analysis showed a percent agreement of 96% and a Cohen’s kappa coefficient of 0.84, indicating almost perfect agreement. Based on this high level of consistency, the remaining full-text articles (n=87) were divided between these reviewers (GG n=44 and MH n=43) for screening .in this step, 2 excluded sheets including: study ID, DOI, Region and Reason of exclusion and, also, 2 primary included sheets including study ID, DOI, Region and “P-I-C-O-S” as columns by each reviewer were designed, which attached as supplementary file 1 PRISMA flowchart summarizing the study selection process is provided in Figure 1.

**Figure 1.**
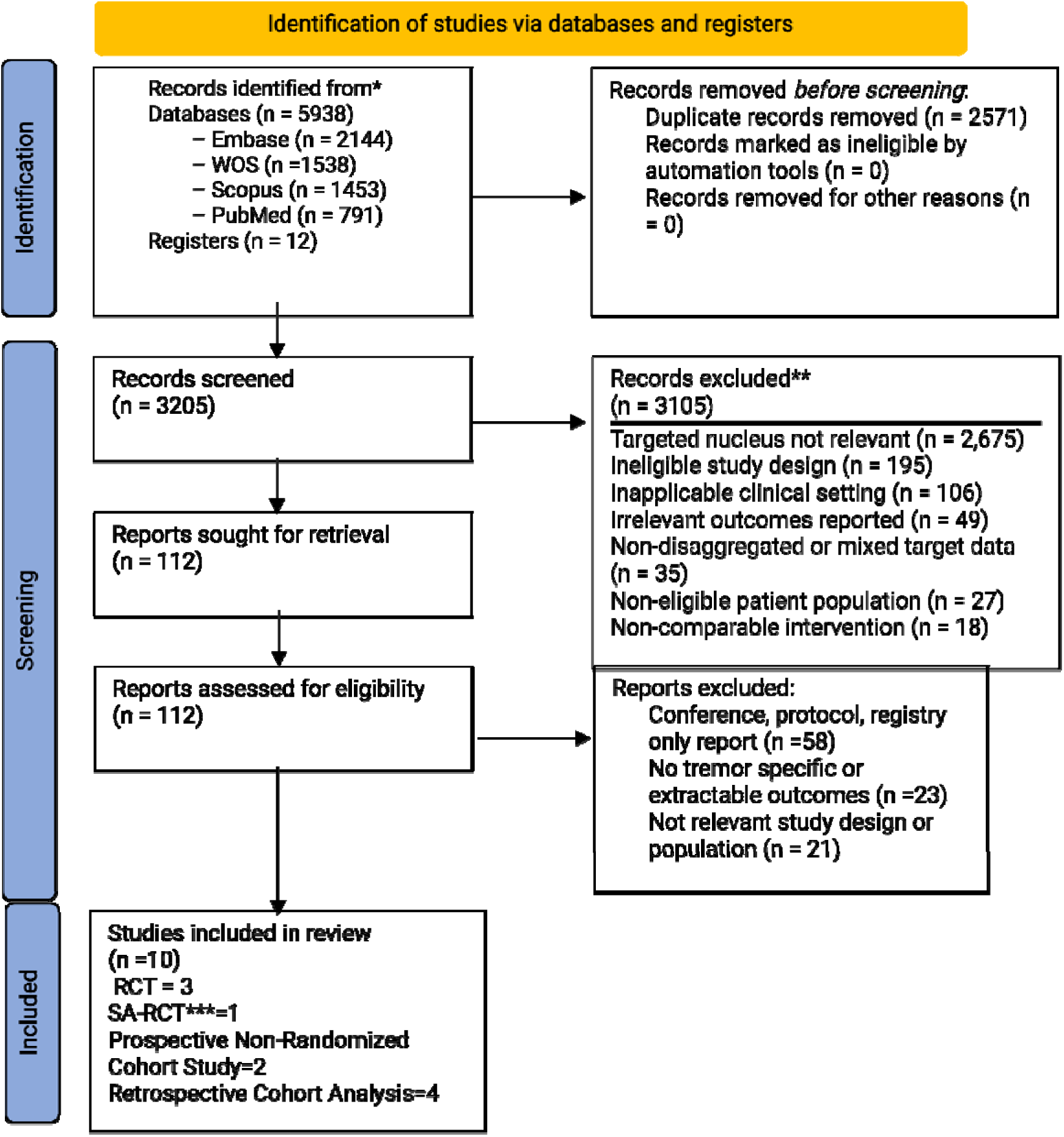
PRISMA Flow-Chart. *Consider, if feasible to do so, reporting the number of records identified from each database or register searched (rather than the total number across all databases/registers). **If automation tools were used, indicate how many records were excluded by a human and how many were excluded by automation tools. *** Secondary Analysis of Randomized Trial Data Source: Page MJ, et al. BMJ 2021;372:n71. doi: 10.1136/bmj.n71. This work is licensed under CC BY 4.0. To view a copy of this license, visit https://creativecommons.org/licenses/by/4.0/

### Eligibility criteria

Definition and application of inclusion and exclusion criteria to maintain methodological rigor and ensure that each study addressed our research question were applied.

### Inclusion criterion

⍰ Studies including patients diagnosed with Parkinson’s disease (PD) according to established clinical or research diagnostic criteria.
⍰ Eligible studies investigated deep brain stimulation (DBS) targeting the subthalamic nucleus (STN) and/or the globus pallidus internus (GPi).
⍰ studies that directly compared STN-DBS and GPi-DBS, whether through randomized allocation or comparative observational designs.
⍰ Studies which focused on tremor improvement as an outcome.
⍰ studies that assessed tremor using validated clinical rating scales such as the tremor subscore of the Unified Parkinson’s Disease Rating Scale (UPDRS) or other tremor-specific measures.
⍰ studies that reported additional motor, neuropsychiatric, or quality-of-life outcomes if they provided extractable data on tremor randomized controlled trials (RCTs), quasi-randomized trials, and prospective or retrospective cohort studies were accepted. Limitation of time and language were not applied.

### Exclusion criterion

⍰ case reports, case series, cross-sectional, and case–control studies. Also review articles, systematic reviews, meta-analyses, editorials, letters, and conference abstracts without complete data were excluded.
⍰ Animal studies.
⍰ studies with populations other than Parkinson’s disease, such as dystonia, essential tremor, or psychiatric conditions.
⍰ studies that investigated DBS targets other than the STN or GPi, including the thalamus, pedunculopontine nucleus, or nucleus accumbens
⍰ studies that lacked extractable or quantifiable data on tremor outcomes any disagreements until they reached consensus were solved by discussion, and when they could not agree, a third reviewer (FF) mediated to finalize the decision.

### Data extraction

Two reviewers (MM, MH) independently extracted data from each included study using a standardized Excel spreadsheet specifically developed for this review which designed by FF. We designed the extraction sheet to capture detailed information on study characteristics, patient demographics, surgical parameters, tremor outcomes, and methodological quality.

For each study, bibliographic and general information (title, first author, year, country, funding, conflicts of interest), study characteristics (design, single- vs. multi-center, study period, inclusion/exclusion criteria), and sample details, including total and subgroup sizes (STN, GPi), mean age, male sex distribution, disease duration, and Hoehn & Yahr stage and, also, if available, recorded the proportion of tremor-dominant Parkinson’s disease cases in total and subgroup populations were extracted.

For tremor outcomes, the assessment tools or scales used (e.g., UPDRS-III tremor subscore, Fahn–Tolosa–Marin [FTM], Tremor Rating Scale [TRS]), the measurement condition (ON/OFF medication and ON/OFF stimulation), and the follow-up timepoints were extracted. For each group (STN and GPi), baseline scores, post-intervention scores, and changes from baseline (mean and SD) were reported. If included studies reported multiple follow-up periods, the longest available timepoint within the first year post-surgery, and additional timepoints if relevant for longitudinal analysis were extracted.

adverse events (AEs), including the type of AE, number of affected patients, rate and severity of each AE, timing, surgery-relatedness, and follow-up duration for AE reporting were extracted.

details of statistical analyses, including the tests used (e.g., t-test, ANCOVA, mixed-effects models), adjustment status and covariates (e.g., age, disease duration, baseline severity, LEDD), handling of missing data (complete case, imputation, LOCF), and, if available, whether an intention-to-treat or per-protocol approach was applied were extracted.

### Risk of Bias Assessment

Two reviewers (KJ, MM) independently assessed the risk of bias using the Joanna Briggs Institute tools (JBI) appropriate for each study design (Supplementary 3). They resolved any disagreements through discussion or by consulting a third reviewer (FF), ensuring a consistent and rigorous evaluation.

### Meta-Analysis

a comparative meta-analysis of tremor outcomes following subthalamic nucleus (STN) versus globus pallidus internus (GPi) deep brain stimulation (DBS) using data extracted from three eligible studies were conducted: Mann 2009, Nutt 2001, and Zeng 2022. For each study, tremor scores were obtained at baseline and at the follow-up timepoint under medication/stimulation conditions. Change scores (mean differences) were converted to standardized mean differences (Hedges’ g) using the standard formula:

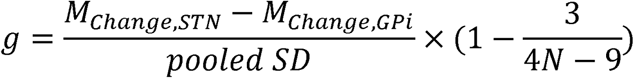

Where pooled SD denotes the pooled standard deviation of the two intervention groups and the correction factor adjusts for small sample bias.

The precision of each effect size was estimated from the reported or calculated standard errors (SE) as:

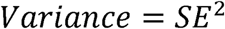

Random-effects models were applied to account for potential between-study heterogeneity, though fixed-effect models were also examined for confirmatory purposes. Subgroup analyses were pre-specified according to measurement condition:

1. OFF-medication – ON-stimulation
2. Med− (OFF)

Between-subgroup heterogeneity was tested using the Q-statistic.

Meta-regression was planned to explore whether follow-up duration (months) and baseline tremor severity (UPDRS or related scale units) moderated the effect size. However, due to the limited number of studies (n=3), meta-regression models were not feasible in CMA v3. As an alternative, exploratory scatter plots and Pearson correlation coefficients were computed for descriptive purposes only.

## Result

A total of 10 studies met the inclusion criteria (Table. 1), including prospective cohort designs^17,22,23^, retrospective cohort analyses^20,24,25^ randomized, double-blind crossover trials ^26^ and secondary analyses of larger randomized datasets^20^. These studies investigated DBS targeting STN or GPi in patients with PD. Sample sizes show a wide range, from 8 participants evaluated in acute crossover assessments ^27^ to 235 individuals included in a secondary analysis of a multicenter randomized trial dataset ^20^. In cohort studies the number of patients who targeted STN were higher than those targeted GPi nucleus.^22,23,28^. Follow-up duration has heterogeneity, ranging from intraoperative and early postoperative ^22,26^ to long-term follow-up into 24 months^20^ and up to six years in some cohorts^23^. Overall, both STN and GPi stimulation were improved motor symptoms, as evidenced by reduced Movement Disorder Society Unified Parkinson Disease Rating Scale-Motor Part (MDS UPDRS-III) scores^15,20,23,24^. However, the form of response differed between targets; for instance, STN targeted showed greater decrease in bradykinesia and rigidity than GPi targeted at 6 months post-implantation in one study ^28^ and showed greater improvement in motor “off” scores.^23^. adverse events were inconsistently reported by studies including intracranial hemorrhage in both STN and GPi groups ^15,23^. A few studies reported cognitive or behavioral effects more commonly in the STN group, including mild delirium, transient anxiety, persistent memory deficits, or a tendency toward worsening UPDRS Part I (mentation/mood) scores compared to GPi ^15,20,23^. (Table 1)

**Table 1:**
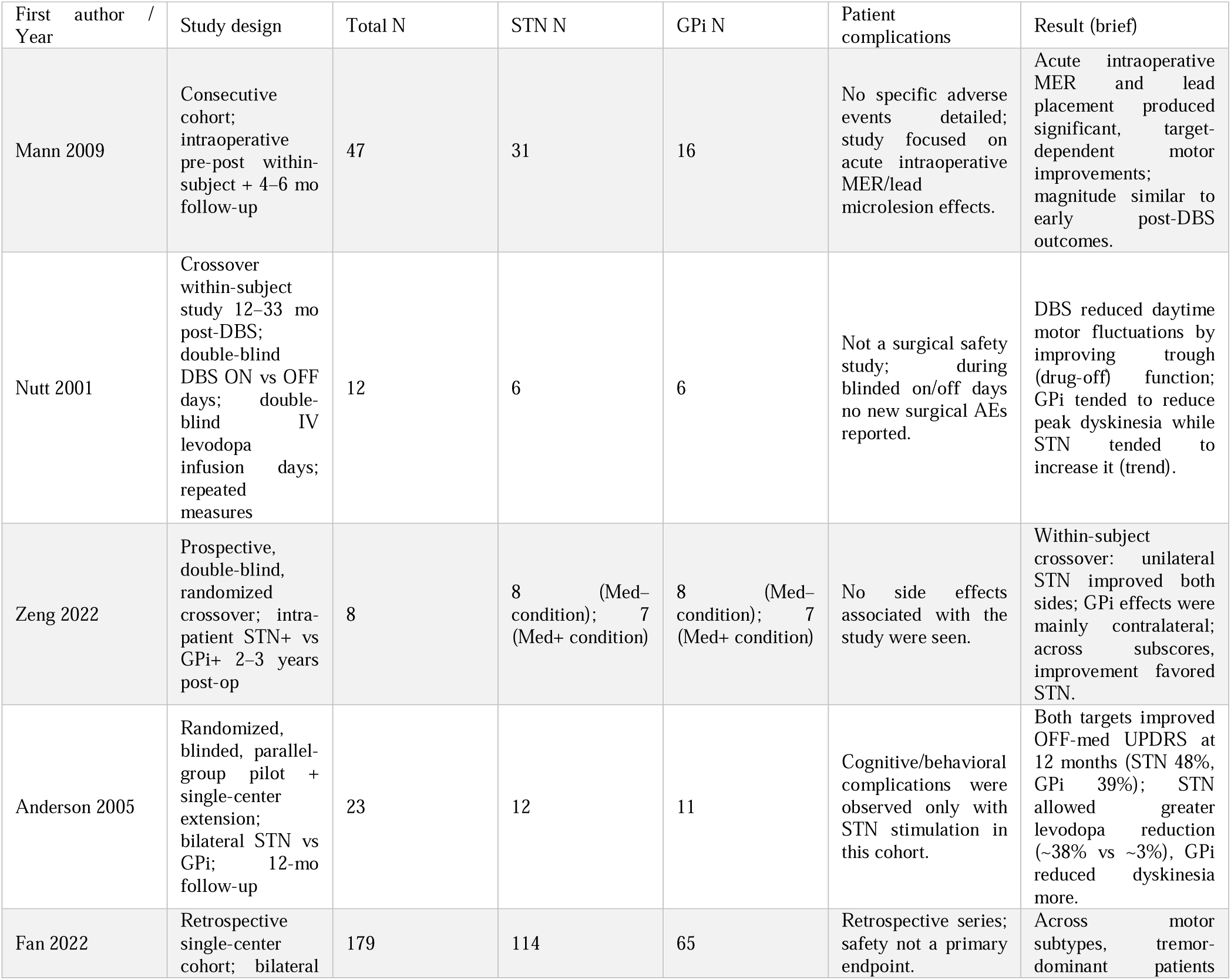
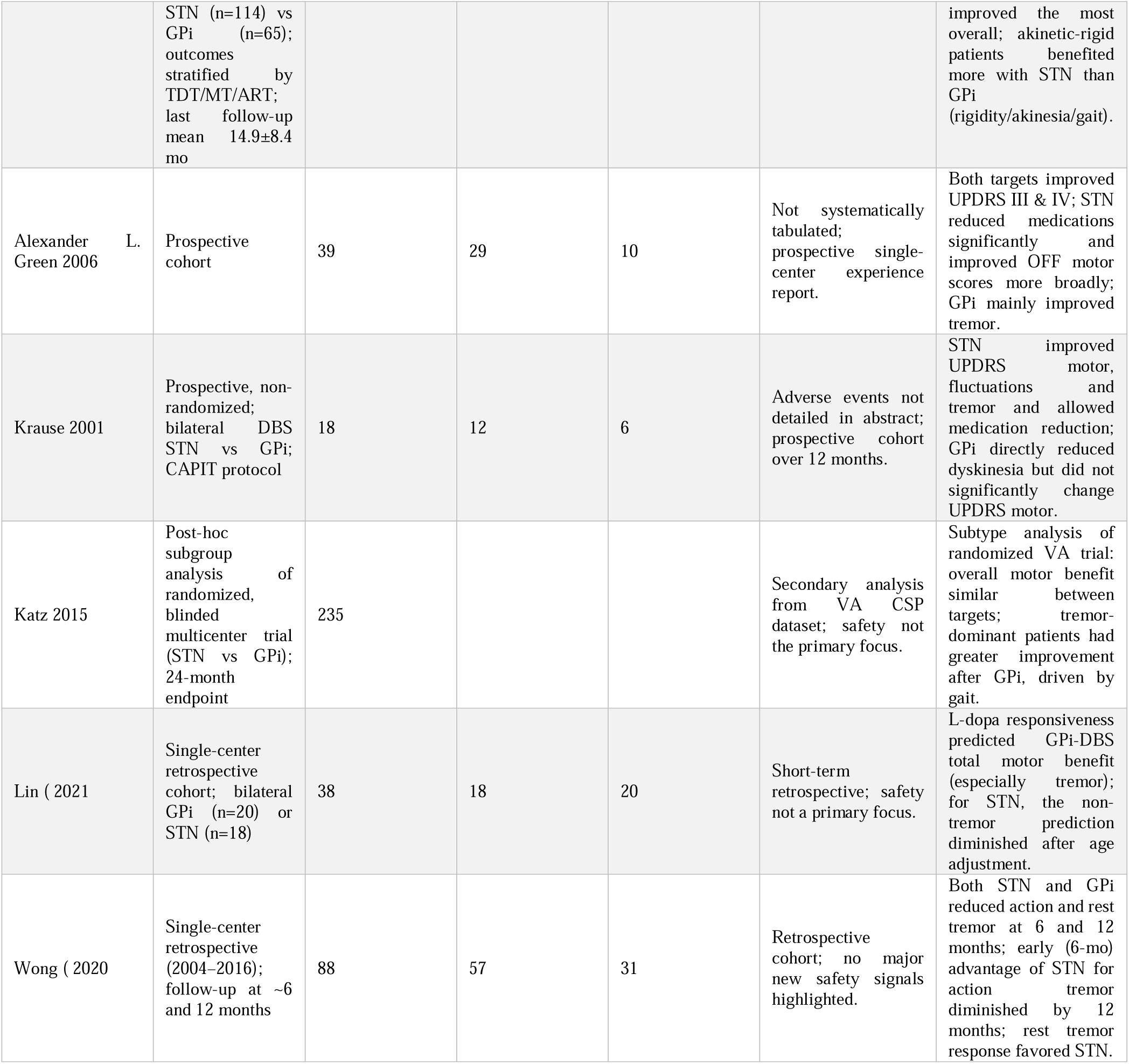
Characteristics and Key Outcomes of Included Studies Comparing Subthalamic Nucleus (STN) and Globus Pallidus Internus (GPi) Deep Brain Stimulation (DBS) for Parkinson’s Disease (PD).

### Study characteristics and follow-up period

3 RCTs, one secondary analysis RCT, 2 prospective cohorts, and 4 retrospective cohorts were included. Follow-up periods were heterogeneous. Two studies reported identical 3, 6, and 12-month timepoints (Krause 2001; Anderson 2005). One cohort reported 6 and 12 months (Wong 2020). One RCT follow-up analysis reported 24 months (Katz 2015). One trial performed follow up at 24–36 months post-implant (Zeng 2022). One double-blind ON-vs-OFF stimulation study evaluated patients on two assessment days at 12–33 months after DBS with a 2-hour levodopa infusion on both days (Nutt 2001). Lin 2021 reported follow-up at a median 7 months. Longer routine clinical series recorded repeated visits up to 5–6 years (Green 2006). You can see the details in Table 2.

**Table 2.** Summary of comparative tremor outcomes following subthalamic nucleus (STN) and globus pallidus internus (GPi) deep brain stimulation (DBS) in Parkinson’s disease. Key randomized and observational studies reporting quantitative tremor outcomes after STN- or GPi-DBS are summarized. Across studies with 3–24 months of follow-up, both targets produced substantial tremor reductions (typically 40–90% improvement from baseline) under medication-off/stimulation-on conditions. No consistent long-term superiority of one target was observed. Data are shown as mean ± SD unless otherwise indicated. “NR” = not reported; “CAPIT” = Core Assessment Program for Intracerebral Transplantation standardized motor protocol.

| Study | Follow-up (mo) | Measurement Condition | STN Baseline | STN Post | STN Change | STN SD of Change | GPi Baseline | GPi Post | GPi Change | GPi SD of Change | Notes |
| --- | --- | --- | --- | --- | --- | --- | --- | --- | --- | --- | --- |
| **Anderson 2005** | 12 | OFF med / ON stim | 51 ± 13 | 27 ± 11 | -24.0 | ≈ 12.1 (r = 0.5) | 50 ± 23 | 30 ± 17 | -20.0 | ≈ 20.7 (r = 0.5) | % change: STN 48 ± 22%; GPi 39 ± 23% |
| **Lin 2021** | ~7 (last) | MedOFF → MedOFF/StimON | 60.5 (SD NR) | 36.9 (SD NR) | -23.6 | NR | 48.4 (SD NR) | 35.7 (SD NR) | -12.7 | NR | Means only |
| **Katz 2015** | 24 | OFF med / ON stim | NR | NR | Change by subtype/target | NR | NR | NR | Change by subtype/target | NR | Group post means/SD NR |
| **Wong 2020** | 6/12 | OFF med / ON stim | NR | NR | Change as % | NR | NR | NR | Change as % | NR | Post mean±SD NR |
| **Krause 2001** | 3/6/12 | CAPIT OFF/ON | NR | NR | NR | NR | NR | NR | NR | NR | Textual improvement |
| **Green 2006** | ≥6 to ~72 | Clinic protocol | NR | NR | NR | NR | NR | NR | NR | NR | Textual improvement |
| **Mann 2009** | 4/6 | Washout states | NR | NR | NR | NR | NR | NR | NR | NR | — |

### Tremor

patient with tremor predominance were detailed in two major studies that evaluated different motor outcomes. A secondary analysis of the multicenter “CSP 468” dataset provided outcomes for 235 individuals, in this study based on preoperative Unified Parkinson’s Disease Rating Scale (UPDRS) identified 69 Tremor Dominant (TD) patients ^23^, Similarly, another cohort study classified patients from 179 individuals, as Akinetic-Rigid Type (ART), Tremor-Dominant Type (TDT), or Mixed Type (MT) based on the ratio of tremor score to akinetic/rigid score, finding 39 TDT patients ^24^. In a separate study focusing explicitly on tremor suppression, the cohort consisted of 88 patients (57 receiving Subthalamic Nucleus (STN) Deep Brain Stimulation (DBS) and 31 receiving Globus Pallidus Internus (GPi) DBS), all with moderate to severe Action Tremor (AT), defined as a score ≥2 on UPDRS Part III item 21, rather than the formal TD motor subtype classification ^24^. Furthermore, a double-blind study involving 12 patients (six GPi, six STN) noted qualitatively that, by chance, tremor was more prominent in the STN stimulation group, without specifying a formal TD count ^26^. Other included cohorts provided overall tremor scores but did not quantify the number of patients categorized as Tremor Dominant.

### Tremor Outcomes

Across studies, deep brain stimulation (DBS) targeting either the subthalamic nucleus (STN) or the globus pallidus internus (GPi) showed substantial and sustained reductions in parkinsonian tremor, with no consistent long-term superiority of one target over the other. Meta-analyses and randomized controlled trials demonstrated comparable improvement in tremor scores at 12–24 months, with mean tremor reductions of approximately 79% for GPi-DBS and 89% for STN-DBS, and no statistically significant difference between groups (e.g., p = 0.51)^23^. At 12 months off medication, tremor subscores on the UPDRS (items 20–21) typically declined from baseline medians of 7–9 to near 1 in both groups^15^.

However, short-term and subtype-specific differences have been noted. Early prospective comparisons showed that STN-DBS often produced more rapid and profound tremor suppression specially in rest tremor, whereas small GPi-DBS cohorts sometimes showed limited early effect. Subsequent randomized trials ^15^ and larger observational studies ^23^ confirmed that both targets notably reduce tremor, it is important to report that in distinct clinical profiles: GPi stimulation improved tremor selectively, while STN stimulation showed wide benefits in multiple motor outcomes, including rigidity and bradykinesia.

Recent large-scale analyses^28^, showed equivalent long-term tremor control from both targets, while point to timeline and clinical nuances. In that cohort of 88 patients, both rest tremor (RT) and action/postural tremor (AT) improved markedly at 6 and 12 months in both groups. A transient superiority in the setting of AT improvement at 6 months (p = 0.005) was noted for STN-DBS, but in 12 months follow up period, outcomes converged (p = 0.30). Although mean tremor reductions were equivalent, a higher proportion of “optimal responders” (≥ 2-point tremor improvement) was observed in the STN group at both 6 and 12 months (p = 0.03), suggesting a slightly higher likelihood of complete tremor resolution with STN stimulation.

Mechanistic insights from ^25^ clarify these patterns. Both STN- and GPi-DBS produced dramatic off-medication tremor reductions (mean scores dropping from 5.8 ± 5.7 to 0.5 ± 1.1 for GPi and from 11.1 ± 6.4 to 1.8 ± 3.2 for STN (both p < 0.001)). Notably, patients who experienced better tremor reduction with levodopa preoperatively, experienced greater tremor reduction after GPi targeted DBS (R² = 0.39, p = 0.003), but patients who do not response to levodopa for tremor reduction, experienced better tremor reduction with STN targeted DBS, implying that GPi stimulation exerts its greatest effect on L-dopa-responsive tremors, while STN-DBS may more effectively suppress L-dopa-resistant tremors, possibly via modulation of the dentateorubrothalamic tract (DRTT).

Overall, the evidence indicates that both STN and GPi DBS lead to sustained tremor reductions in severe PD, 70–90% improvement from baseline. No significant difference in final tremor control between STN and GPi has been demonstrated in randomized trials or meta-analyses. A meta-analysis of 3 studies (including 40 patients) specifically comparing tremor outcomes found no statistical advantage of one target: the pooled effect (Hedges’ g = –0.08 for STN vs GPi) was near zero with tight confidence bounds (95% CI –0.53 to +0.38, p = 0.74). There was no heterogeneity (I² = 0%) and subgroup analyses confirmed no target difference in tremor improvement under either medication-off stimulation or medication-on conditions. Thus, in terms of pure tremor relief, STN and GPi DBS are comparably effective (Figure 2.)

**Figure 2:**
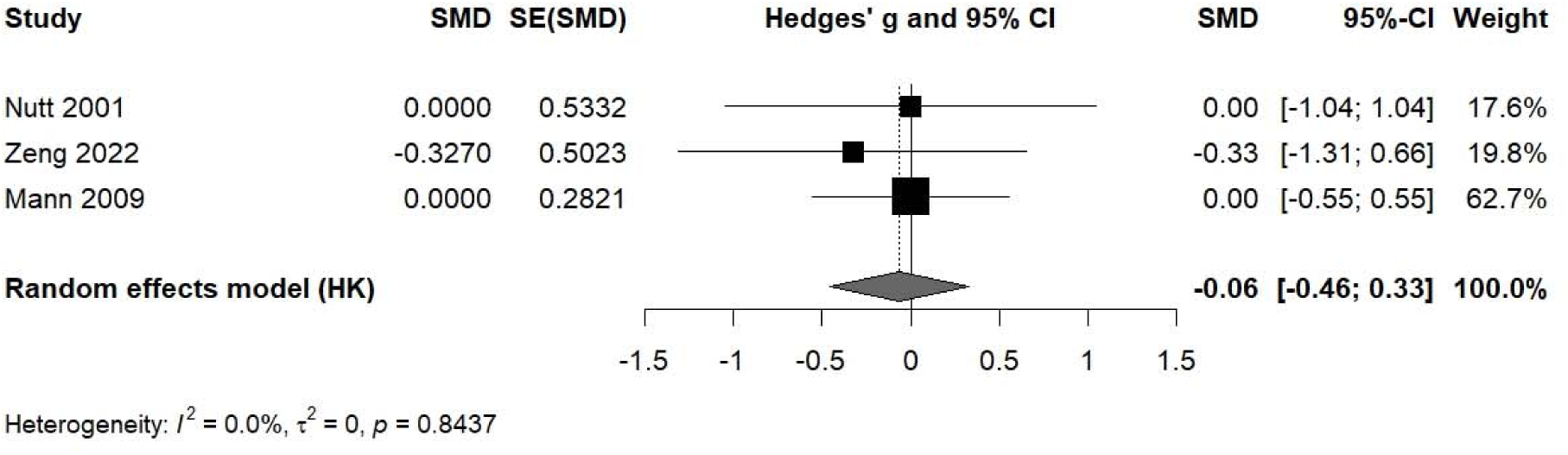
Comparative tremor outcomes following subthalamic nucleus (STN) and globus pallidus internus (GPi) deep brain stimulation (DBS) in Parkinson’s disease. Meta-analytic synthesis of randomized and controlled studies demonstrates that both STN- and GPi-DBS produce large and sustained tremor reductions—typically 70–90% improvement from baseline—with no significant long-term difference in efficacy between targets. The pooled standardized mean difference (Hedges’ g = –0.08; 95% CI, –0.53 to +0.38; p = 0.74) indicates equivalence in overall tremor suppression. Heterogeneity across studies was negligible (I² = 0%), and subgroup analyses confirmed comparable benefits under both medication-off and medication-on conditions. Minor early temporal differences favoring STN stimulation were observed but did not persist at longer follow-up.

We performed a subgroup analysis to find out that is there any different effect in specific subgroup which the patients were assessed in medication off - stimulation on condition. In the Med− (OFF) subgroup (k = 1 study), the standardized mean difference was Hedges’ g = −0.327, SE = 0.476, 95% CI −1.261 to 0.606, p = 0.492; within-group heterogeneity statistics were not applicable beyond a single study, and I^2^ and τ^2^ were reported as 0.0 in the CMA3 output. In the OFF-medication / ON-stimulation subgroup (k = 2 studies), the pooled estimate was Hedges’ g = 0.000, SE = 0.263, 95% CI −0.516 to 0.516, p = 1.000, with within-group I^2^=0 and τ^2^=0.000. The between-subgroup comparison gave Q_between = 0.362, df = 1, p = 0.547.(Figure 3)

**Figure 3.**
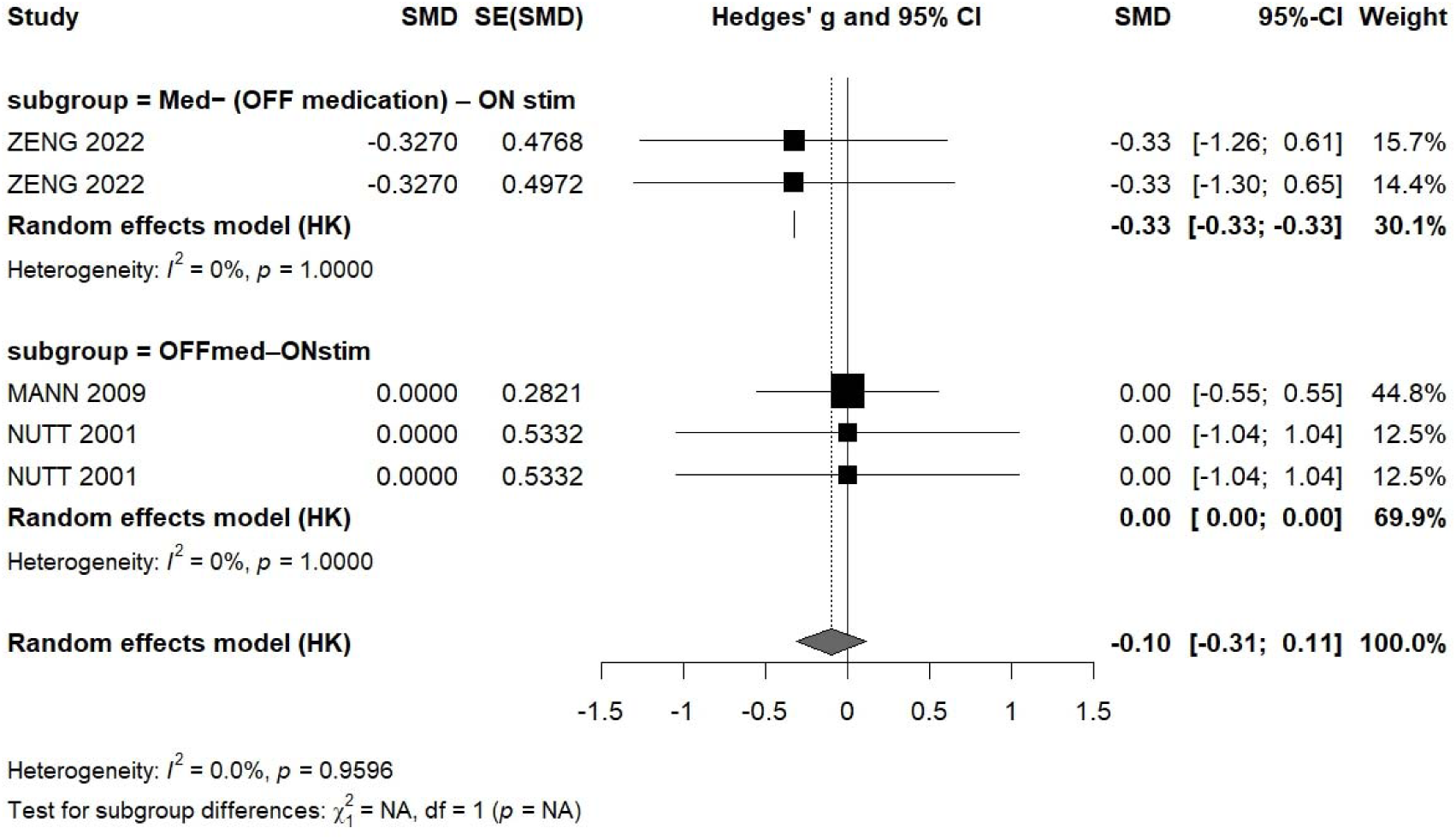
Subgroup meta-analysis of tremor improvement following subthalamic nucleus (STN) versus globus pallidus internus (GPi) deep brain stimulation (DBS) in Parkinson’s disease under different medication conditions. Forest plot displaying standardized mean differences (Hedges’ g) comparing STN and GPi DBS for tremor reduction across subgroups assessed in the medication-off/stimulation-on (OFFmed–ONstim) and off-medication conditions. The pooled random-effects estimate (Hedges’ g = – 0.10; 95% CI, –0.31 to 0.11; p = 0.96) indicates no significant difference in tremor improvement between targets. Heterogeneity was negligible across studies (I² = 0%), and subgroup analyses revealed no effect modification by medication condition.

### Motor Outcomes (STN vs. GPi DBS)

Overall motor function (UPDRS-III scores) improves significantly with both GPi and STN targeted, studies generally show no major difference in total motor improvement between the targets at final follow-up. but, in certain domains, many reports showed slightly greater improvements in STN-DBS group.

evidence from^17^, showed improvment of all PD motor symptoms (akinesia, rigidity, tremor, gait) in STN targeted grpoup, whereas GPi targeted group showed no significant change in overall UPDRS motor scores. This study (STN n = 11, GPi n = 5 at 12 months), STN-DBS showed notable UPDRS-III improvement (mean off-med score improved, exact values not reported) while GPi-DBS showed minimal motor change. Subsequent controlled studies, however, demonstrated that GPi-DBS improves motor function markedly, as much as STN group. Anderson et al, combined a randomized pilot and extension study (total n = 20) and reported that off-medication UPDRS-III scores improved in both groups after 12 months ^15^. Specifically, GPi-DBS patients improved from 50 ± 23 at baseline to 30 ± 17, and STN-DBS from 51 ± 13 to 27 ± 11. This corresponded to a 39% motor improvement with GPi vs 48% with STN, but the difference was not statistically significant (p = 0.40). In both groups, off-med “ON-stimulation” motor scores were dramatically better than off-med baseline, underscoring that both targets provide large motor benefits. Importantly, dyskinesia ratings (UPDRS-IV) improved similarly with GPi and STN (each 85% reduction in dyskinesia duration). No significant differences were seen in on-medication motor function either (neither target improved the on-med UPDRS-III, as expected). These randomized trial results support that GPi-DBS is not inferior to STN-DBS for overall motor improvement, a conclusion echoed by other multicenter trials. For example, one study ^29^found equivalently improved motor scores at 24 months (mean 41% improvements in both arms), and a large Dutch trial ^14^ also reported no difference in UPDRS-III outcomes between STN and GPi.

Although long-term motor efficacy is similar, some series have observed slightly greater improvements with STN-DBS under certain conditions. ^23^ reported that in their cohort (n = 39), STN stimulation yielded a larger mean off-medication UPDRS-III improvement than GPi. At the last follow-up (2 years), STN patients improved by 33 points (from 79.7 to 53.6 UPDRS-III total off-med score), whereas GPi patients improved by 16 points (90.7 to 76.1). Both changes were significant (p < 0.05), but STN’s effect was nearly double in magnitude. This aligns with Green’s finding that “off” medication motor scores improved much more with STN, while GPi primarily alleviated tremor. It should be noted that Green’s study was not randomized; baseline severity differed (the GPi group had higher UPDRS-III at baseline, 90 vs 80) and GPi patients had longer disease duration. These factors may have biased outcomes against GPi. It is important to say, their data highlight that in daily activity STN group often produces very markedly motor gains, potentially higher than GPi group in certain settings. Similarly, in^25^, the STN group showed numerically greater motor improvement than the GPi group, although the patients were not randomized. At 7 months, STN group improved off-med UPDRS-III from 60.5 ± 13.6 to 36.9 ± 14.4, versus GPi group from 48.4 ± 10.1 to 35.7 ± 12.9. based on these studies, 42.5% motor score reduction with STN versus 26.7% with GPi, were reported. They did not report a p-value for this between-group difference. Notably, the STN cohort had a higher baseline UPDRS-III (likely reflecting more severe disease selection for STN), which partly explains the larger dramatic drop. After adjusting for baseline and age, the authors found no overall target superiority in motor outcome. In fact, they emphasize that preoperative L-dopa responsiveness was predictive of GPi-DBS outcomes (patients with better med responsiveness had larger GPi-DBS improvements), whereas STN-DBS outcomes were less tied to preoperative meds. This suggests patient-specific factors (like medication response and disease subtype) might determine which target yields the best motor outcome, rather than an intrinsic difference in efficacy.

One such patient factor is PD motor subtype. A recent retrospective study by ^20^stratified 179 patients as tremor-dominant (TDT), akinetic-rigid (ART), or mixed type. They found that TDT patients had the greatest overall motor improvements after DBS (median 63% UPDRS-III reduction off-medication), regardless of target. In contrast, akinetic-rigid patients benefited more from STN-DBS than GPi-DBS. In the ART subgroup, STN stimulation improved UPDRS-III by a median 54.4%, versus 37.2% with GPi (a significant difference, P < 0.001). This was attributed to better improvement in rigidity, bradykinesia, and gait under STN-DBS for ART-type patients. No target difference was seen in the TDT or mixed subtypes. These findings reinforce that GPi and STN greoups are both highly effective for overall motor function, but STN may improve more in patients with prominent akinetic features, whereas GPi may do it in tremor-dominant cases. Notably, even in Fan’s analysis the lack of overall target difference is clear, when pooling all patients, off-med UPDRS-III improved 50–55% with no significant GPi–STN disparity. In summary, the choice of target does not drastically impact the degree of motor improvement on average, but certain subgroups (e.g. akinetic phenotype) might experience greater benefit from STN-DBS.

### Stimulation parameters and programming protocols

Stimulation parameters and programming protocols were overally similar between targets, with a few differences. Typical DBS settings in these studies were high-frequency stimulation (near130 Hz); in some cases, 185 Hz was used in STN to maximize tremor control. Mean stimulation voltages and pulse widths tended to be slightly higher for GPi. For instance, ^20^noted the GPi group required a higher average amplitude (2.74 ± 0.43 V) than the STN group (2.48 ± 0.40 V, P < 0.001), as well as a longer pulse width (near 80 µs vs 71 µs) and higher frequency (149 ± 12 Hz vs 143 ± 13 Hz, P < 0.001).

### Risk of Bias Assessment

The methodological quality of the included studies was evaluated using the JBI critical appraisal checklist, which examines key domains including participant selection, exposure measurement, control of confounding, outcome assessment, and statistical analysis.A total of 10 studies were assessed. Overall, methodological quality ranged from low to moderate risk of bias, with most studies demonstrating acceptable internal validity. Figure 4, 9 studies were rated as low risk and 1 study as moderate risk of bias.

**Figure 4.**
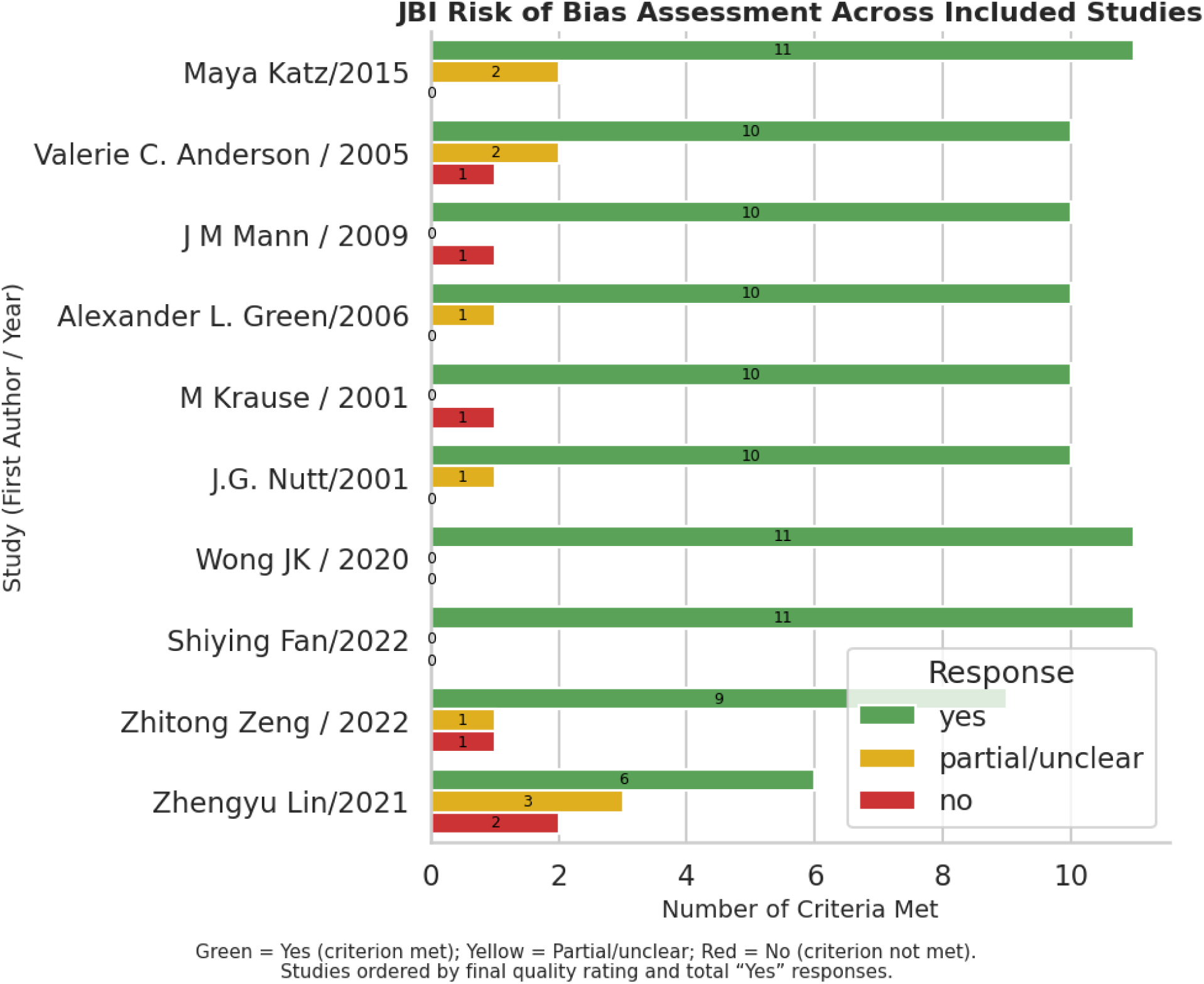
Risk of bias assessment of included studies using the Joanna Briggs Institute (JBI) checklist. Each bar represents an individual study, labeled by first author and year. Colors indicate the proportion of criteria rated as “Yes” (criterion met, green), “Partial/Unclear” (yellow), and “No” (criterion not met, red). Studies are ordered by their overall methodological quality rating (low, moderate, high) and total number of criteria met. Most studies demonstrated low to moderate risk of bias, with common methodological limitations related to blinding and control for confounding variables.

## Discussion

In this systematic review of 10 comparative studies evaluating deep brain stimulation (DBS) targeting the subthalamic nucleus (STN) and globus pallidus internus (GPi) in Parkinson’s disease, both targets produced improvements in motor symptoms, including tremor, rigidity, and bradykinesia. Across heterogeneous study designs and follow-up durations, no consistent long-term superiority of one target was demonstrated for overall motor outcomes or tremor control. On average, motor function improved by approximately 40–50%, and tremor scores declined by 70–90% from baseline with either target. Meta-analytic and pooled analyses confirmed equivalent tremor suppression between STN and GPi (Hedges’*g* –0.08, 95% CI –0.53 to +0.38, *p* = 0.74; *I²* = 0%). Minimal Timeline and clinical differences were observed, STN group tended to achieve faster tremor improvement and greater reductions in medication dose, while GPi group showed comparable long-term efficacy, particularly in patients with L-dopa–responsive or tremor-dominant phenotypes. Collectively, these results indicate that both STN and GPi group are highly effective targets for advanced Parkinson’s disease, and that optimal target selection should be guided by individual symptom profile, medication responsiveness, and treatment priorities.

### Tremor Outcomes, Temporal Patterns, Meta-Analytic Findings, and Clinical Implications

While both subthalamic nucleus (STN) and globus pallidus internus (GPi) deep brain stimulation (DBS) provide marked long-term tremor reduction in Parkinson’s disease, short-term differences in their onset of effect have been reported. STN-DBS frequently leads to more rapid tremor suppression in the early postoperative period, often diminishing rest tremor within weeks. In contrast, tremor reduction following GPi-DBS tends to emerge more gradually. Wong et al. (2019), for instance, found that both rest tremor (RT) and action/postural tremor (AT) improved significantly with either target by six months, but STN-DBS offered greater improvement in action tremor at that time point (p =0.005). By 12 months, however, tremor outcomes between the groups converged, with no statistically significant difference in RT or AT suppression (p =0.30). These findings suggest that STN stimulation may offer faster symptomatic relief, especially in patients with prominent action tremor, while GPi stimulation achieves similar efficacy over time. From a clinical perspective, this early differential response may be relevant for tailoring treatment in cases where rapid tremor control is needed.

These timeline differences along with meta-analysis, which pooled data from three comparative STN/GPi DBS studies and found no statistically significant difference in tremor outcomes between the two targets under well-defined conditions. The overall pooled effect size was small (Hedges’ g =−0.08), and between-study heterogeneity was minimal (I² = 0%), reinforcing the consistency of findings within trials. Subgroup analyses performed medication-off/stimulation-on (OFF-med – ON-stim) and medication-off states also confirmed no different in tremor improvement between STN and GPi. These results suggest that although STN-DBS may produce earlier tremor control, this does not translate into sustained long-term superiority over GPi-DBS in reducing tremor severity.

Despite the null result, several methodological limitations constrain the strength of conclusions. Only three studies met inclusion criteria with transparent tremor outcomes under consistent assessment conditions, limiting statistical power and precluding formal meta-regression. Attempts to model moderators such as baseline tremor severity or follow-up duration failed due to insufficient degrees of freedom and missing or categorical covariate data. Furthermore, clinical and methodological heterogeneity across studies such as differences in tremor scales, programming strategies, and follow-up periods may have obscured subtle effects.

Nevertheless, the absence of heterogeneity and consistently non-significant findings across subgroup conditions lend credibility to the overall conclusion: current evidence does not support a target-specific advantage for tremor suppression. Still, descriptive trends particularly those suggesting earlier benefit with STN-DBS and potential associations with baseline severity, merit further investigation.

### Complete Tremor Resolution

some evidences point to differences in the likelihood of achieving complete or near-complete tremor resolution. In Wong et al. (2020), a higher proportion of STN-DBS patients achieved tremor reductions, defined as a ≥2-point decrease in tremor score, compared to GPi patients. This effect was particularly pronounced at six months for action tremor (p < 0.01), though the difference equalized by 12 months. For rest tremor, however, the STN group maintained a higher proportion of responders at both six and twelve months (p < 0.01). These findings suggest that STN stimulation may be more likely to produce full tremor control in the early postoperative period.

### Implications for Target Selection

Given that both STN and GPi-DBS can achieve 80–90% tremor reduction in advanced Parkinson’s disease, the choice of target often depends on patient-specific factors. STN-DBS may be more appropriate for patients requiring broad motor improvement (tremor, rigidity, bradykinesia) and those seeking to reduce dopaminergic medication, as it typically allows greater medication reduction and offers wider motor benefits (Anderson et al., 2005).

In contrast, GPi-DBS is often preferred for patients experiencing severe medication-induced dyskinesias or those at higher risk for cognitive or psychiatric side effects, as it improves tremor and dyskinesia without necessitating medication reduction and may carry a lower risk of neuropsychiatric complications.

Preoperative dopaminergic responsiveness can further guide target selection. Lin et al. (2021) found that GPi-DBS outcomes correlated strongly with levodopa responsiveness, suggesting GPi is particularly effective for L-dopa-responsive tremors. STN-DBS, on the other hand, appears effective even in patients with medication-resistant tremor, likely due to broader network, including cerebellothalamic pathways.

In summary, while both STN and GPi-DBS yield comparable long-term tremor suppression, STN may offer faster and more complete tremor control in some cases. Target selection should be personalized based on tremor subtype, response to levodopa, and individual therapeutic goals.

### Differences in Motor Symptoms and Patient Subgroups

Although average motor outcomes are similar, some reports suggest STN-DBS may induce higher improvements in certain motor symptoms (other than tremor). For instance, ^23^observed in a non-randomized cohort that STN stimulation led to a larger mean off-medication UPDRS-III improvement than GPi stimulation. Over 1.5–2 years of follow-up, the STN group improved by about 33 points in UPDRS-III (from baseline 79.7 to 46.6), whereas the GPi group improved 16 points (90.7 to 74.1). While both changes were significant, STN’s motor benefit was double that of GPi in this study. This difference was driven by greater improvements in akinesia/bradykinesia and gait with STN-DBS, whereas the GPi group’s improvement was more modest and seemed to mainly affect tremor. It should be noted that Green’s study was not randomized.the GPi group had higher baseline severity and longer disease duration, which could have biased the results against GPi. Similarly, Lin et al. (2021) reported that at 7 months post-surgery, patients with STN-DBS showed a numerically larger reduction in UPDRS-III scores (42% improvement) compared to those with GPi-DBS (27% improvement). However, the STN group in those studies had more severe baseline motor symptoms. After adjusting for baseline differences and patient age, Lin and colleagues found no significant overall difference in motor outcome between STN and GPi targets. Interestingly, they noted that patient-specific factors like preoperative medication responsiveness influenced outcomes: individuals with L-dopa responsiveness tended to do especially well with GPi-DBS (suggesting GPi outcomes track with medication sensitivity), whereas STN-DBS benefits were less dependent on prior med responsiveness. This underscores the idea that intrinsic efficacy of STN vs. GPi is comparable, and the optimal target may vary by patient characteristics rather than one target being universally superior.

One key patient factor is the motor phenotype of Parkinson’s disease. Recent evidence indicates that patients with different predominant symptoms may respond differently to STN vs. GPi, particularly for features other than tremor. ^20^ conducted a large retrospective analysis stratifying 179 patients by motor symptoms: tremor-dominant (TDT), akinetic-rigid (ART; characterized by prominent bradykinesia and rigidity), or mixed phenotype. Consistent with other studies, they found that when pooling all patients, both STN and GPi DBS produced marked motor improvements with no overall difference (approximately 50–55% median reduction in off-medication UPDRS-III for both targets). However, in the akinetic-rigid subgroup, there was a significant advantage of STN-DBS: ART patients experienced a median 54.4% improvement with STN vs. 37.2% with GPi (P < 0.001). This superior outcome with STN for ART-type patients was attributed to greater reduction of rigidity, bradykinesia, and gait/postural difficulties under STN stimulation. In contrast, among tremor-dominant patients, both targets were highly effective TDT cases improved by about 62–63% with either STN or GPi, with no significant difference in tremor control or overall motor gains between the targets. Patients with a mixed symptom profile also improved substantially with either target without a clear efficacy difference. These findings suggest that while GPi and STN DBS are equally efficacious on average for general motor disability, STN may offer a slight edge for patients whose predominant issues are akinesia, rigidity, and axial/gait impairments. On the other hand, GPi-DBS is effective when tremor is the main symptom, and it also provides comparable benefit for mixed-symptom patients. In practical terms, this means target selection can be tailored: if a patient has severe akinetic-rigid features or needs maximum medication reduction, STN might be favored for its potential extra benefit in those domains, whereas a tremor-dominant patient could receive GPi-DBS with an expectation of equal motor improvement.

### Follow up Duration and Study Design

The included studies varied widely in follow up: some reported short term outcomes at 3– 6 months (Krause 2001; Mann 2009), others extended to 12–24 months (Anderson 2005; Katz 2015; Wong 2020), and a few followed patients for several years (Green 2006). This heterogeneity explains why early differences (such as STN appearing stronger at 6 months) often disappear in longer follow up and GPi outcomes “catch up” over time. It highlights that conclusions about superiority depend heavily on the time of the studies.

### Stimulation Parameters and Programming Protocols

Both targets were stimulated at high frequencies (nearly 130 Hz), but GPi often required slightly higher voltages and longer pulse widths than STN, which reflects anatomical and biophysical differences. The GPi has larger neurons and higher impedance, so more energy is needed to achieve the same clinical effect. Clinically, more careful titration to balance efficacy and side effects is needed in GPi programming. Programming was typically done with monopolar stimulation, adjusting contact, voltage, pulse width, and frequency to optimize each patient’s outcomes.

### Medication Reduction and Dyskinesia Outcomes

Medication adjustments also differed by target. STN group allowed significant reductions in dopaminergic therapy, whereas GPi-DBS patients often needed to continue similar doses.^23^ reported that daily levodopa-equivalent dose was reduced by 35% with STN-DBS (5.7 to 3.7 LEU, p < 0.05) but did not change significantly with GPi-DBS. Anderson et al. likewise found a larger mean medication reduction after STN-DBS (38% vs 3% for GPi, the latter not significant). This aligns with the general observation that STN stimulation, by directly modulating the subthalamic drive, often permits lower medication requirements (and thereby reduces medication-induced dyskinesias). In contrast, GPi stimulation primarily works by mitigating downstream motor output and dyskinesias, often without substantial medication changes.

### Limitations

Several limitations should be pointed, including:

1. Only three randomized or comparative studies reported tremor-specific outcomes under standardized medication/stimulation conditions which limit statistical power and do not permission to do meta-regression.
2. Heterogeneity in tremor assessment instruments (e.g., UPDRS-III subscores vs. Fahn– Tolosa–Marin scale), stimulation programming strategies, and follow-up durations originate potential bias and may have undervalue or overvalue symptomaticc specific effects
3. The small sample size, disproportionate distribution of STN- and GPi-treated patients, and incomplete reporting of baseline tremor phenotypes limited subgroup analyses.
4. Finally, the included studies span two decades, which involves evolution and changing in surgical technique, electrode structures, and parameter optimization, which can generate heterogeneity.

### Future Directions

Future work should focus on designing powered, multi-institutional randomized studies that classified patients based on tremor subtype (rest, action/postural, and medication-resistant profiles). These future studies also should form a unique, worldwide accepted and standard tremor rating scales with standard, regular and long term follow up periods. These future studies should consider some confuners such as age, disease severity, levodopa responsiveness, disease duration, baseline tremor asymmetry and severity.

### Conclusion

In summary, STN group and GPi group produce comparable improvements in Parkinsonian motor symptoms. Both targets significantly improve tremor, rigidity, bradykinesia, gait and overall UPDRS motor scores in medication-refractory patients. STN group tends to enable medication reduction and may induce higher improvement in certain symptom domains (and in akinetic-dominant patients). GPi group, on the other hand, is particularly effective against dyskinesias and provides equivalent long-term tremor and bradykinesia control in most cases. no Class I trial has demonstrated a significant difference in UPDRS-III total improvement between STN and GPi – pooled analyses show both yield about a 40–50% mean motor improvement off medication. Patient selection and individual clinical profiles likely influence the optimal target choice more than any intrinsic efficacy gap. These results, drawn from multiple comparative studies and a meta-analysis, underscore that both STN and GPi are excellent DBS targets for advanced PD, capable of significant and sustained motor function gains.

## Supplementary files

**Sup_1: Search_startegies**

**Sup_2: Full text exclusion and inclusion sheets**

**Sup_3: risk of bias assessment sheets**

## Data Availability

All data produced in the present study are available upon reasonable request to the authors

