## Supplementary material for "Subthalamic Nucleus(STN) versus Globus Pallidus Internus(GPi) targeted with Deep Brain Stimulation in Parkinson’s Disease: A Systematic Review and Meta-Analysis of tremor outcome": Sup_1

**-Pubmed search strategy :**

**( "Parkinson Disease"[Mesh] OR parkinson*[tiab] OR PD[tiab] ) AND ( "Deep Brain Stimulation"[Mesh] OR "deep brain stimulation"[tiab] OR DBS[tiab] ) AND ( ("Subthalamic Nucleus"[Mesh] OR "subthalamic nucleus"[tiab] OR STN[tiab]) AND ("Globus Pallidus"[Mesh] OR "globus pallidus internus"[tiab] OR "internal globus pallidus"[tiab] OR GPi[tiab]) ) AND ( compar*[tiab] OR head-to-head[tiab] OR versus[tiab] OR vs[tiab] OR random*[tiab] OR trial[tiab] )**

**-Scopus search strategy:**

**( TITLE-ABS-KEY ( parkinson* OR PD ) )**

**AND ( TITLE-ABS-KEY ( "deep brain stimulation" OR DBS ) )**

**AND ( TITLE-ABS-KEY ( "subthalamic nucleus" OR STN )**

**AND TITLE-ABS-KEY ( "globus pallidus internus" OR "internal globus pallidus" OR GPi ) )**

**AND ( TITLE-ABS-KEY ( compar* OR "head-to-head" OR versus OR vs OR random* OR trial ) )**

**-Web of science Search strategy :**

**TS=(parkinson* OR PD)**

**AND TS=("deep brain stimulation" OR DBS)**

**AND ( TS=("subthalamic nucleus" OR STN)**

**AND TS=("globus pallidus internus" OR "internal globus pallidus" OR GPi) )**

**AND TS=(compar* OR "head-to-head" OR versus OR vs OR random* OR trial)**

**-Embase search strategy:**

**( 'parkinson disease'/exp OR parkinson*:ti,ab OR PD:ti,ab )**

**AND ( 'deep brain stimulation'/exp OR 'deep brain stimulation':ti,ab OR DBS:ti,ab )**

**AND ( ('subthalamic nucleus'/exp OR 'subthalamic nucleus':ti,ab OR STN:ti,ab)**

**AND ('globus pallidus'/exp OR 'globus pallidus internus':ti,ab OR 'internal globus pallidus':ti,ab OR GPi:ti,ab) )**

**AND ( compar*:ti,ab OR 'head-to-head':ti,ab OR versus:ti,ab OR vs:ti,ab OR random*:ti,ab OR trial:ti,ab )**
